# Improving Neonatal Nurses’ Knowledge and Preventive Practices for Unplanned Extubation: Effective of a low-cost Multimodal Educational Intervention in the NICU

**DOI:** 10.64898/2026.10.08.26365033

**Authors:** Reem Ahmad Ali, Ayat Wael Al Hourani, Islam Salameh Al-Aseat

## Abstract

Unplanned Extubation is a frequent adverse event in Neonatal Intensive Care Units (NICUs), that is associated with increased morbidity, prolonged hospitalization and financial costs. Evidence suggests that unplanned extubation (UE) is preventable through the management of its risk factors. Many modifiable UE risk factors are directly related to nursing care, thus empowering nurses through education can improve neonatal safety outcomes. This study aimed to evaluate the effectiveness of a structured educational intervention in improving nurses’ knowledge and practices related to UE in NICUs. A quasi-experimental pretest-posttest study was conducted in two tertiary hospitals in northern Jordan. One hospital is university-affiliated and one is governmental hospital. Participating Nurses in the selected settings received a four-week multimodal educational intervention related to UE. The intervention comprised of face-face session, WhatsApp-based educational messages, educational videos and placement of a poster in the NICU. Knowledge and practices of 58 nurses were assessed at baseline, post intervention and one month later using a validated questionnaire and direct observation. A baseline rate of UE was also measured in both settings with rate of 5 events per 100 ventilator days in the university-affiliated compared to 32 events per 100 ventilator days in governmental hospital. Repeated-measures and mixed-design ANOVA revealed significant improvements in nurses’ knowledge in both settings with a large effect size (ηp² = .60). Also, the observed preventive practices scores also improved significantly and remained elevated at one-month follow-up (ηp² = .50). Older age was associated with lower baseline knowledge, though this was diminished post study intervention. Whereas no demographic variables were associated with practice scores. The high rates of UE highlight the need for quality improvement. Study findings suggest that UE-focused education is a feasible and cost-effective strategy for strengthening neonate nurses’ competencies regarding safer care for mechanically ventilated neonates in the NICUs.

## Introduction

Unplanned extubation (UE) is the unintended removal of an endotracheal tube (ETT) regardless of the underlying reason for its displacement or removal (1). In neonatal intensive care units (NICUs), UE represents a significant patient safety concern because it may result in acute complications, including oxygen desaturation, bradycardia, the need for resuscitation (2), reintubation, increased risk of infection (3) and prolonged hospitalization (4,5). UE has also been associated with longer-term morbidities, such as bronchopulmonary dysplasia and acquired subglottic stenosis (6,7). Reported UE rates vary considerably across NICUs, ranging from 0.42 to 7.2 events per 100 ventilator days (2,8–13). Such variation may partly reflect differences in UE definitions and reporting methods. Although no national data on UE rates have been officially published in Jordan, the event remains an important concern given its preventable nature and potential consequences for neonatal outcomes.

Multiple factors contribute to UE in the NICU and can be broadly categorized as people-related, environment-related, and procedure- and measurement-related factors (14–18). Patient-related factors include low gestational age, low birth weight, agitation, and excessive movement, whereas provider- and parent-related factors include inconsistent adherence to preventive practices and inadequate support during handling activities. Environmental factors encompass workload, staffing patterns, and the availability of clear protocols. Procedure- and measurement-related factors include ETT securement, positioning, suctioning, repositioning of infants, monitoring practices, event reporting systems, and communication among healthcare providers. Parental handling and kangaroo care have also been identified as potential risk situations when appropriate education and support are lacking (19).

Because nursing care intersects with many of the modifiable risk factors associated with UE, neonatal nurses are central to prevention efforts. Nurses provide continuous bedside surveillance of mechanically ventilated neonates and are directly involved in ETT securement and retaping, airway suctioning, positioning, routine handling, monitoring tube placement, and assisting with diagnostic and therapeutic procedures (14). In addition, nurses coordinate care among multidisciplinary team members and provide education and support to families. Consequently, improving nurses’ knowledge and adherence to evidence-based preventive practices may contribute substantially to reducing UE and improving neonatal safety outcomes. Although several UE risk factors are nonmodifiable at the individual nurse level, educational interventions can target nursing behaviors and decision-making processes that influence many modifiable procedural and monitoring-related risk factors.

Evidence indicates that UE rates can be reduced through adherence to standardized prevention bundles that include consistent ETT securement methods, appropriate ETT positioning, a two-person approach during high-risk procedures, and standardized handling practices (8,9,16,17,20). Educational initiatives, simulation training, staff workshops, and competency-based programs have been associated with improved compliance with preventive practices and reductions in UE incidence (8,9,12,16,17,21,22). Furthermore, staff education has been identified as a key component of effective UE prevention strategies (23,24).

Although comprehensive UE prevention bundles have demonstrated effectiveness, their implementation often requires substantial organizational resources, multidisciplinary coordination, continuous auditing, and ongoing performance monitoring. Such requirements may present challenges in resource-constrained settings. Educational interventions, in contrast, offer a feasible and cost-effective strategy for promoting adherence to evidence-based practices while targeting several modifiable risk factors associated with UE. Therefore, due to financial constraints and limited access to participating clinical settings, the present study focused specifically on staff education as a practical and scalable intervention for improving UE prevention practices among neonatal nurses.

Despite the growing body of research on UE, a significant evidence gap remains, as most studies evaluate practice compliance and UE incidence without concurrently assessing nurses’ knowledge and preventive practices. Given the pivotal role of nurses in UE prevention, research addressing this gap is warranted to provide evidence that can guide educational strategies, practice improvement initiatives and neonate safety outcomes in the NICU.

Accordingly, this study aimed to evaluate the effectiveness of a structured educational intervention in improving neonatal nurses’ knowledge and nursing practices within the observed NICU. Although UE incidence was assessed prior to the implementation of study intervention as a contextual patient-safety indicator, the primary outcomes were nurses’ knowledge and preventive practices. By focusing on staff education as a feasible and scalable intervention, this study sought to address an important evidence gap and support safer care for mechanically ventilated neonates.

## Materials and Methods

### Study Design

A quasi-experimental pretest-posttest study design was used to evaluate the effectiveness of an educational intervention in improving neonatal nurses’ knowledge and practices related to the prevention of unplanned extubation (UE) in neonatal intensive care units (NICUs). Outcomes were assessed at baseline, one week after the intervention, and one-month post-intervention to evaluate both immediate and sustained effects.

### Study Setting

The study was conducted in the NICUs of the two largest tertiary referral teaching hospitals in northern Jordan. Both hospitals serve as major referral centers with high neonatal admission rates. The university-affiliated NICU (U-NICU) contains 31 incubators, employs 53 nurses, and operates with an average nurse-to-neonate ratio of 1:3. The governmental NICU (G-NICU) comprises 46 incubators, employs 48 nurses, and has an average nurse-to-neonate ratio of 1:5.

### Participants and Sampling

Registered nurses providing direct neonatal care in the NICU for at least one year were eligible to participate. Additional inclusion criteria included holding at least a bachelor’s degree in nursing, proficiency in English, and provision of voluntary informed consent.

Eligible nurses were approached face-to-face by a member of the research team and informed about the study objectives, procedures, confidentiality measures, voluntary participation, and right to withdraw at any time without penalty. All study participants provided written informed consent. Ethical approval was obtained from the Institutional Review Board (No. 17/163/2023).

A convenience sampling strategy was employed. Sample size was estimated using G*Power 3.1 based on a moderate effect size (0.50), α = 0.05, and power = 0.80, yielding a minimum required sample of 34 participants. To maximize statistical power, all eligible nurses from both NICUs were invited to participate. Of the 95 eligible nurses, 58 completed the study (Fig. 1), including 36 nurses from G-NICU and 22 from U-NICU. Reasons for nonparticipation included competing work commitments, lack of interest in research participation, and research fatigue among some nurses who had recently participated in other studies. All enrolled participants completed the study, resulting in no participant attrition. [Fig. 1 Enrollment in the Study]

**Figure 1.**
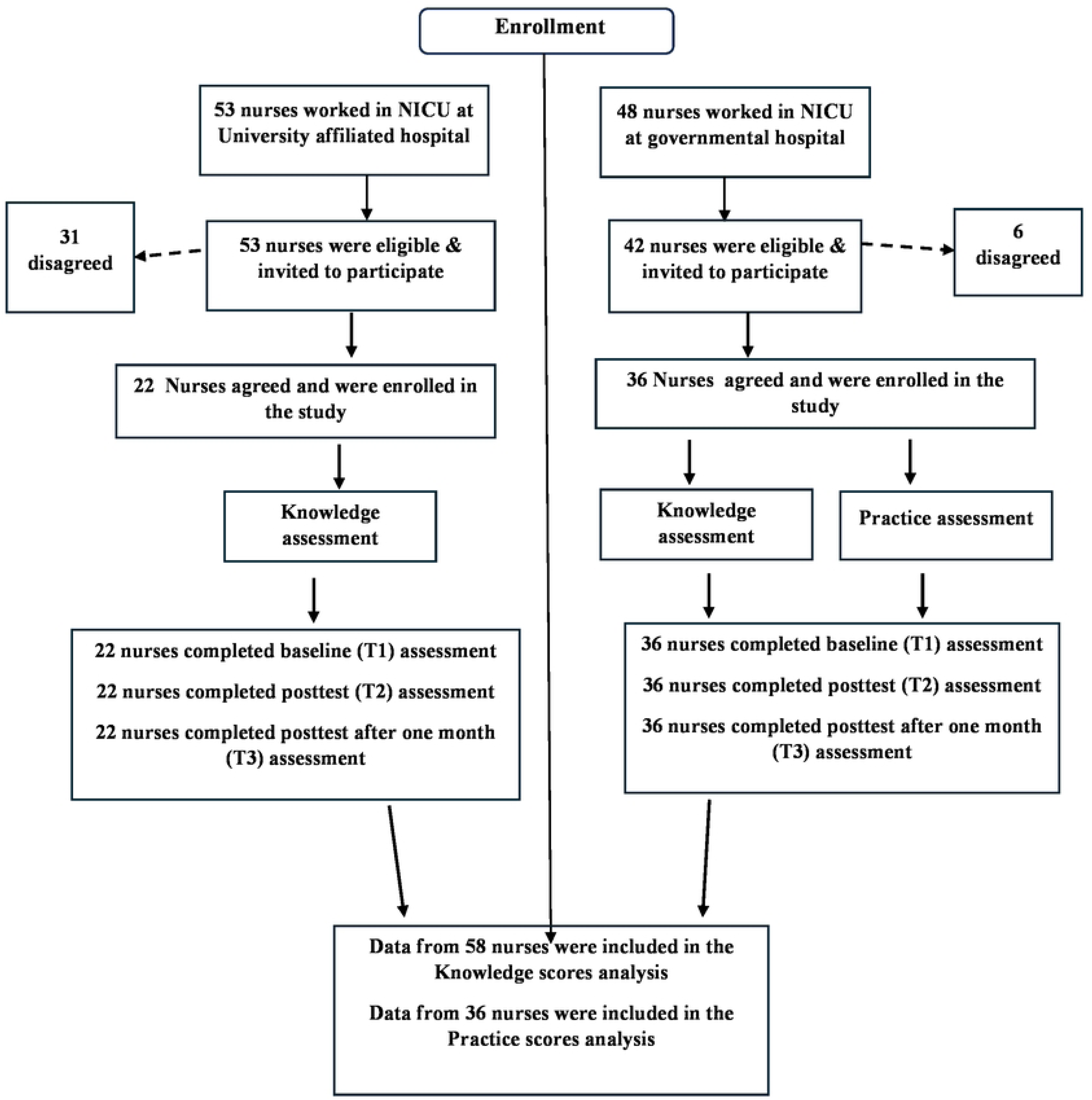
Enrollment in the study.

### Educational Intervention

The educational intervention was developed from current evidence related to UE prevention and neonatal airway management (3–6,14–17,25–33) and relevant international guidelines including *Actionable Patient Safety Solutions: Unplanned Extubation* (34) and neonatal intubation guidance (35).

The intervention content was reviewed and approved by an expert panel. It was delivered over four weeks using a multimodal approach consisting of WhatsApp-based educational materials, instructional videos, interactive face-to-face sessions, and educational posters summarizing UE risk factors and evidence-based preventive practices. Content focused on UE epidemiology and consequences, neonatal ETT management, suctioning practices, securing and positioning of intubated neonates, UE risk factors, and evidence-based prevention strategies. The interactive sessions were delivered by the same researcher for both study settings. Additionally, identical educational posters were displaced in both settings. A summary of intervention components is presented in S1 File.

### Study Measures

A structured questionnaire and observational checklist were used to assess nurses’ knowledge and practices related to UE prevention. Scale items for knowledge scale and observational checklist were developed from the evidence-based recommendations and preventive measures incorporated into the educational intervention. Both study intervention and developed instruments were informed by the same literature sources. All instruments were administered in English, which is the primary language of nursing education in Jordan.

#### Knowledge of Unplanned Extubation

Nurses’ knowledge was assessed using self-reported questionnaire. The knowledge scale comprised 58 items across five domains; including Characteristics and epidemiology of UE, Risk factors for UE, complication, neonatal endotracheal intubation and UE prevention measures. Items were presented as multiple-choice or true/false/don’t know questions. Correct answers received one point, while incorrect and "don’t know" responses received zero points. Higher scores indicated greater knowledge of UE and its prevention. Complete knowledge items are provided in S2 File.

#### Nursing Practices Related to UE Prevention

Nursing practices were evaluated using a structured observational checklist. The checklist contained 15 items assessing adherence to recommended practices for preventing UE, including respiratory assessment, suctioning techniques, ventilator tubing management, and maintenance of head and neck alignment during handling and repositioning. Each item was scored as correctly performed (1) or not performed/incorrectly performed (0), with higher scores indicating greater adherence to recommended practices. Complete observational checklist items are provided in S3 File.

Because certain practices were governed by institutional policies rather than individual nurses, organizational-level variables were assessed separately. These included ETT securement methods, staffing requirements during radiographic procedures, repositioning, tape changes, and parental handling of intubated neonates. Due to logistical and access constraints, direct observation of practice was limited to G-NICU; therefore, practice-related findings should be interpreted as reflective of this setting only.

#### Unplanned Extubation Rate

Prior to implementation of the intervention, UE incidence was prospectively monitored and expressed as the number of UE events per 100 ventilator days. Data were collected using a standardized monitoring form that recorded daily numbers of intubated neonates, ventilator days, and UE events requiring reintubation. Forms were completed daily by the nurse in charge at each participating NICUs. This standardized metric allowed comparison of UE rates between units.

### Validity, Reliability, and Pilot Testing

Face and content validity of the questionnaire, observational checklist, and educational materials were evaluated by a multidisciplinary expert panel comprising neonatologists, respiratory therapists, critical care nursing specialists, and a nursing professor. The content validity index was 0.90, indicating excellent content validity.

Internal consistency of the knowledge questionnaire was excellent (Cronbach’s α = 0.916), while the observational checklist demonstrated acceptable reliability (Cronbach’s α = 0.77).

A pilot study was conducted to assess the clarity, feasibility, and comprehensibility of the study instruments and procedures. Five nurses from U-NICU completed the questionnaire, and four nurses from G-NICU participated in pilot observations before implementation of the intervention. The pilot findings indicated that the instruments were clear, relevant, and feasible for use in the clinical setting. As no modifications were required, pilot data were retained and included in the final analysis.

### Data Collection Procedures

Data collection was conducted between 1^st^ of March and 30^th^ of December 2024. As exhibited in figure 2 following baseline monitoring of UE rates, nurses’ practices were observed using the study checklist. Owing to logistical constraints, practice observations were conducted only in G-NICU. [Fig 2. Flow chart of the study protocol]

**Figure 2.**
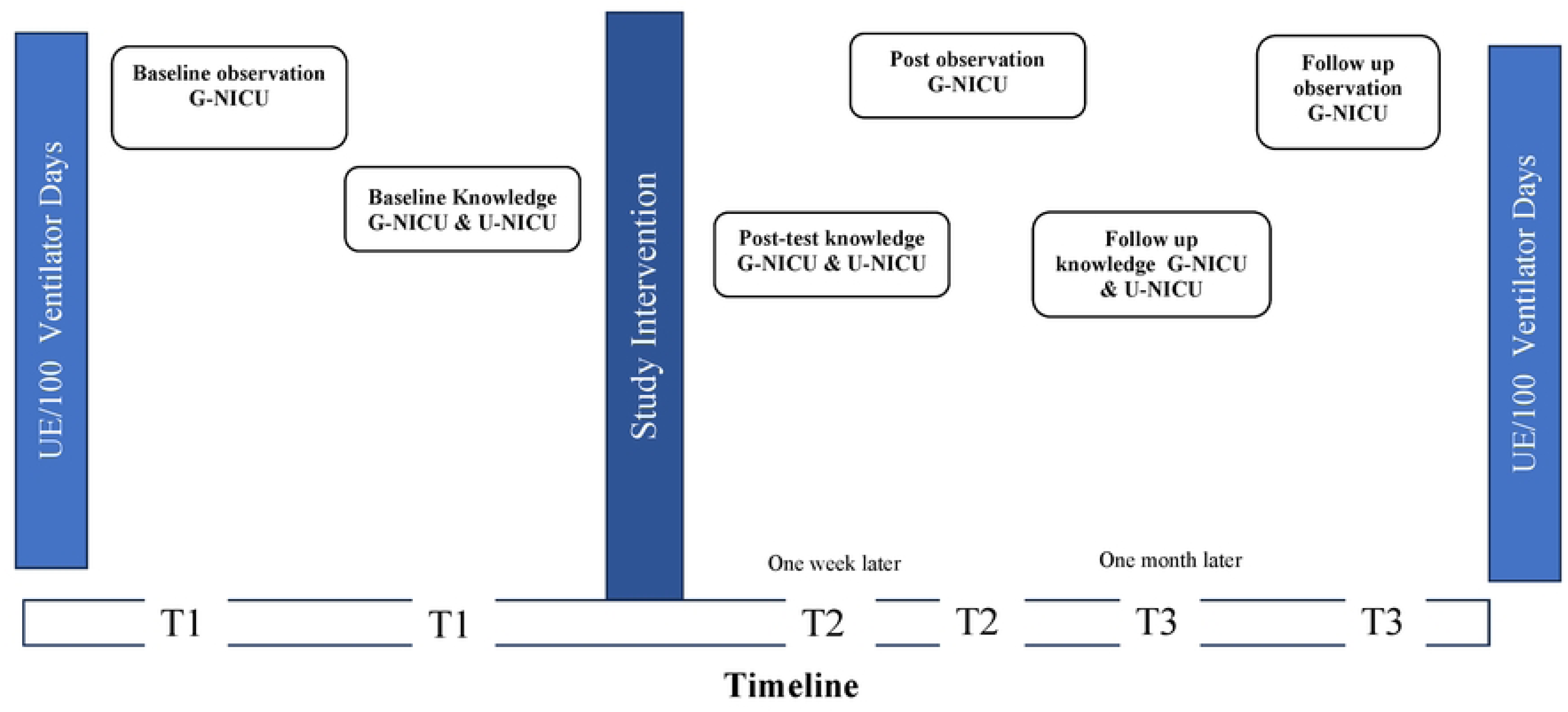
Flow chart of the study protocol.

Intubated neonates were assigned to participating nurses according to routine staffing procedures. Observations were conducted by a trained member of the research team, following standardized training provided by the principal investigator. Each observation covered an entire nursing shift and focused on routine clinical care rather than emergency situations.

Following completion of baseline observations, nurses completed the knowledge questionnaire via Google Forms. Questionnaire links were distributed through WhatsApp. Knowledge was assessed in both NICUs at baseline, following the intervention, and one month later. Nursing practices were reassessed after the intervention and at follow-up using the same observation procedures and measurement tools.

### Statistical Analysis

Data were analyzed using SPSS version 27. Descriptive statistics were used to summarize participant characteristics and study variables. Independent-samples *t*-tests were conducted to compare outcomes between NICUs. Repeated-measures analysis of variance (RM-ANOVA) was used to evaluate within-group changes in knowledge and practice across the three assessment points. A mixed-design repeated-measures ANOVA (split-plot ANOVA) was performed to examine differences in knowledge trajectories between U-NICU and G-NICU over time. Statistical significance was set at *p* < .05.

## Results

### Participant Characteristics and NICU Profiles

A total of 58 neonatal nurses participated in the study, including 22 (37.9%) from the university-affiliated NICU (U-NICU) and 36 (62.1%) from the governmental NICU (G-NICU). As presented in Table 1, most participants (87.9%) held a bachelor’s degree, and the mean age was 36.1 years. No significant differences were observed between settings regarding educational level, years of experience, previous training, or perceptions of the timing and contributing factors of UE. Notably, none of the participants had previously received formal training on endotracheal tube (ETT) management or UE prevention. The only significant demographic difference was age, with nurses in G-NICU being older than those in U-NICU (*p* = 0.019).

**Table 1.** Socio-demographical characteristics of study sample (N=58)

| Variables | Category | Total sample | University Hospital | Governmental Hospital | Test value | P-value |
| --- | --- | --- | --- | --- | --- | --- |
|  |  | N=58 | N=22<br>(37.9%) | N=36<br>(62.1%) |  |  |
|  |  | N (%) | N (%) | N (%) |  |  |
| Education | Bachelor's degree | 51 (87.9) | 19 (86.4) | 32 (88.9) | ---- | 0.540<sup>F</sup> |
|  | Master's degree | 7 (12.1) | 3 (13.6) | 4 (11.1) |  |  |
| Years of experience in NICU | 1-5 years | 18 (31.0) | 11 (50.0) | 10 (27.8) | 3.113 | 0.211<sup>X</sup> |
|  | 6-10 years | 21 (36.2) | 6 (27.3) | 12 (33.3) |  |  |
|  | More than 10 | 19 (32.8) | 5 (22.7) | 14 (38.9) |  |  |
| Training on ETT care for premature infants | No | 58 (100) | 22 (100) | 36(100) | ---- | ---- |
| Education on UE in NICU? | No | 58 (100) | 22 (100) | 36 (100) | --- | --- |
| Timing UE | Day shift | 7 (12.1) | 5 (22.7) | 2 (5.6) | 3.780 | 0.138<sup>FH</sup> |
|  | Night shift | 8 (13.8) | 2 (9.1) | 6 (16.7) |  |  |
|  | Anytime (day or night) | 43 (74.1) | 15 (68.2) | 28(77.8) |  |  |
|  |  | N=58 | N=22<br>(37.9%) | N=36<br>(62.1%) |  |  |
|  |  | N (%) | N (%) | N (%) |  |  |
| Events precede UE | Suctioning | 24 (41.4) | 13 (59.1) | 11 (30.6) | 5.445 | 0.231 <sup>FH</sup> |
|  | Turning/changing position of infants | 19 (32.8) | 6 (27.3) | 13 (36.1) |  |  |
|  | Weighting infants | 4 (6.9) | 0 (0.0) | 4 (11.1) |  |  |
|  | Lifting infants on X-ray plate | 4 (6.9) | 1 (4.5) | 3 (8.3) |  |  |
|  | During infant care | 7 (12.1) | 2 (9.1) | 5 (13.9) |  |  |
| Age / years<br>(Mean ±SD) |  | 36.09±4.66 | 34.27±2.76 | 37.19±5.24 | 2.414 | **0.019** <sup>t</sup> |

*FH: Fisher-Freeman-Halton Exact Test, X: Chi-square test, t: independent t-test, F: Fisher exact test*

Table 2. summarizes the organizational characteristics of the participating NICUs. Both units used uncuffed oral ETTs, reported limited use of sedation except for severely agitated neonates, lacked UE risk-warning systems and standardized ETT securement methods, and relied on a single nurse during radiographic procedures involving intubated neonates. Several differences were identified between settings. In U-NICU, ETT retaping was typically performed by two nurses using a securement approach similar to the oral Pants technique, whereas in G-NICU retaping was generally performed by a single nurse using a non-standardized method. Parents in U-NICU were permitted to hold and care for intubated neonates under nursing supervision, whereas parental handling of intubated neonates was prohibited in G-NICU. Neither unit used a standardized UE reporting system; event documentation was limited to nursing notes in U-NICU and was absent in G-NICU.

**Table 2.**
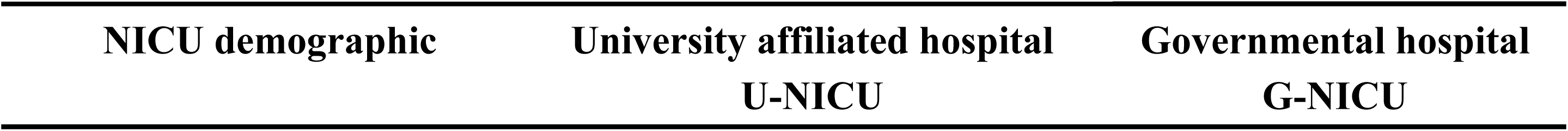
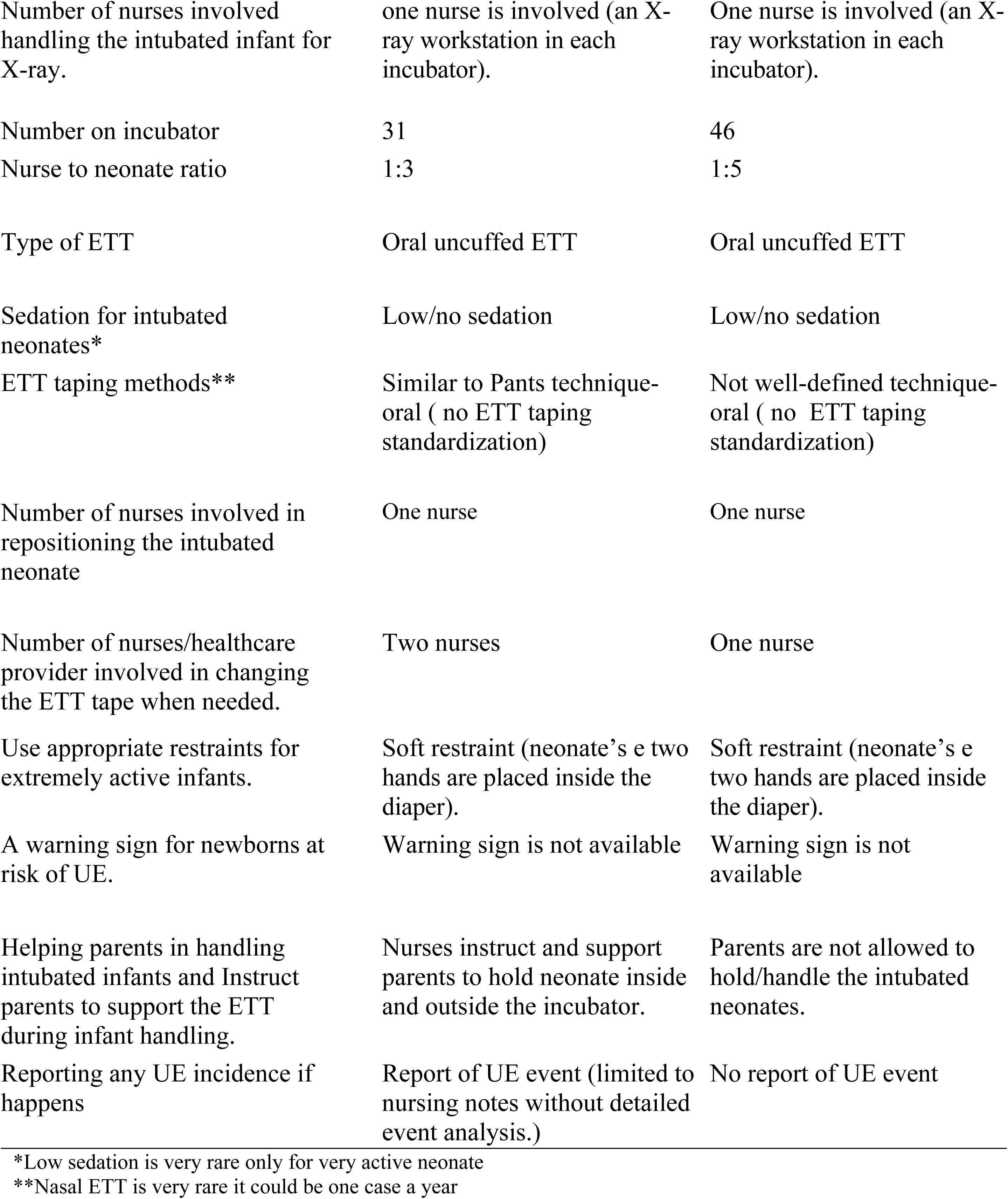
descriptive characteristics of NICU at two settings.

Before implementation of the educational intervention, the UE rate was 5 events per 100 ventilator days in U-NICU and 32 events per 100 ventilator days in G-NICU. Of these events, one and eleven occurred during the morning shift, respectively. The difference was statistically significant (*z* = −4.92, *p* < 0.001; 95% CI: -37.76% to -16.24%), indicating a substantially higher UE burden in G-NICU.

### Effect of the Educational Intervention on Nurses’ Knowledge

Repeated-measures ANOVA demonstrated significant improvements in the nurses’ overall knowledge of UE across the three assessment points in both settings. Improvements occurred primarily between baseline (T1) and immediately after the intervention (T2) and were sustained at the one-month follow-up (T3) (Table. 3)

In U-NICU, significant improvements were observed in knowledge of UE characteristics and epidemiology (*F* = 9.69, *p* = 0.002; ηp² = 0.32), risk factors (*F* = 9.16, *p* < 0.001; ηp² = 0.30), and neonatal endotracheal intubation (*F* = 5.40, *p* = 0.008; ηp² = 0.20). No significant changes were observed in knowledge related to UE complications or preventive strategies.

In G-NICU, significant improvements were observed across all knowledge domains, including UE characteristics and epidemiology (*F* = 133.63, *p* < 0.001; ηp² = 0.79), risk factors (*F* = 100.76, *p* < 0.001; ηp² = 0.74), complications (*F* = 57.01, *p* < 0.001; ηp² = 0.62), neonatal intubation (*F* = 128.78, *p* < 0.001; ηp² = 0.79), and preventive strategies (*F* = 44.75, *p* < 0.001; ηp² = 0.56).

As displayed in Table 4. Split-plot ANOVA analysis of the overall knowledge scores revealed significant effects of time (*F* = 85.25, *p* < 0.001; ηp² = 0.60), hospital setting (*F* = 17.94, *p* < 0.001; ηp² = 0.24), and the time-by-setting interaction (*F* = 21.54, *p* < 0.001, ηp² = 0.28). Although baseline knowledge scores were higher in U-NICU than G-NICU, the difference was not statistically significant (37.36 ± 9.24 vs. 32.33 ± 10.72, *p* = 0.073). Following the intervention, nurses in G-NICU achieved significantly higher knowledge scores than those in U-NICU, both one week after the intervention (56.00 ± 2.04 vs. 45.18 ± 5.92, *p* < 0.001) and at one-month follow-up (53.83 ± 3.84 vs. 44.50 ± 8.39, *p* < 0.001).

**Table 3.** Mean Differences Over Time in Knowledge and Practice Regarding Unplanned Extubation at the Study Settings: Repeated Measures ANOVA Results.

| Settings | NICU at University affiliated hospital (U-NICU)<br>(n= 22) |  |  |  |  |  |  |  |  | NICU at Governmental hospital (G-NICU)<br>(n= 36) |  |  |  |  |  |  |  |
| --- | --- | --- | --- | --- | --- | --- | --- | --- | --- | --- | --- | --- | --- | --- | --- | --- | --- |
| Study outcomes | T1 | T2 | T3 | F-value | P-value | $\eta_p^2$ | Bonferroni test | | | T1 | T2 | T3 | F-value | P-value | $\eta_p^2$ | Bonferroni test | |
|  | Mean (SD) | Mean (SD) | Mean (SD) |  |  |  | Time s | P-value |  | Mean (SD) | Mean (SD) | Mean (SD) |  |  |  | Time s | p-value |
| knowledge of UE |  |  |  |  |  |  |  |  |  |  |  |  |  |  |  |  |  |
| Characteristics and Epidemiology | 3.5<br>(0.96) | 4.86<br>(1.52) | 4.91<br>(1.69) | 9.693 | **0.002** | 0.316 | T1:T2<br>T1:T3<br>T2:T3 | 0.005<br>0.005<br>1.00 |  | 2.92<br>(1.52) | 6.69<br>(0.62) | 6.53<br>(0.77) | 133.63 | **<0.001** | 0.79 | T1:T2<br>T1:T3<br>T2:T3 | <0.001<br><0.001<br>0.886 |
| Risk Factors | 12.82<br>(3.33) | 15.64<br>(2.04) | 15.59<br>(2.79) | 9.164 | **<0.001** | 0.304 | T1:T2<br>T1:T3<br>T2:T3 | 0.003<br>0.026<br>1.00 |  | 11.03<br>(3.60) | 18.19<br>(1.09) | 17.61<br>(1.54) | 100.76 | **<0.001** | 0.74 | T1:T2<br>T1:T3<br>T2:T3 | <0.001<br><0.001<br>0.223 |
| Complications | 5.18<br>(1.87) | 5.91<br>(1.44) | 5.73<br>(1.75) | 1.521 | 0.230 | 0.068 | --- | --- |  | 4.53<br>(2.04) | 7.81<br>(0.52) | 7.08<br>(1.02) | 57.01 | **<0.001** | 0.62 | T1:T2<br>T1:T3<br>T2:T3 | <0.001<br><0.001<br>0.003 |
| Neonatal Intubation | 3.77<br>(1.31) | 5.05<br>(1.33) | 4.77<br>(1.72) | 5.395 | **0.008** | 0.204 | T1:T2<br>T1:T3<br>T2:T3 | 0.012<br>0.102<br>1.00 |  | 3.0<br>(1.89) | 7.61<br>(0.90) | 7.14<br>(1.02) | 128.78 | **<0.001** | 0.79 | T1:T2<br>T1:T3<br>T2:T3 | <0.001<br><0.001<br>0.154 |
| Preventive strategies | 12.09<br>(3.65) | 13.73<br>(1.98) | 13.5<br>(2.32) | 2.617 | 0.085 | 0.111 | --- | --- |  | 10.86<br>(4.16) | 15.69<br>(0.52) | 15.47<br>(0.70) | 44.75 | **<0.001** | 0.56 | T1:T2<br>T1:T3<br>T2:T3 | <0.001<br><0.001<br>0.481 |
| **Overall knowledge** | 37.36<br>(9.24) | 45.18<br>(5.92) | 44.50<br>(8.39) | 7.892 | **<0.001** | 0.275 | T1:T2<br>T1:T3<br>T2:T3 | 0.005<br>0.042<br>1.00 |  | 32.33<br>(10.72) | 56.0<br>(2.04) | 53.83<br>(3.84) | 131.96 | **<0.001** | 0.79 | T1:T2<br>T1:T3<br>T2:T3 | <0.001<br><0.001<br>0.023 |
| **Practice** |  |  |  |  |  |  |  |  |  | 9.44<br>(2.63) | 12.33<br>(1.52) | 12.75<br>(1.41) | 35.41 | **<0.001** | .50 | T1:T2<br>T1:T3<br>T2:T3 | <0.001<br><0.001<br>0.376 |

*T1: baseline, T2 after one week of intervention: T3: post one month. $\eta_p^2$ : partial eta-squared Small 0.01, medium 0.06, large 0.14. Richardson, J.* *T. (2011)*

**Table 4.**
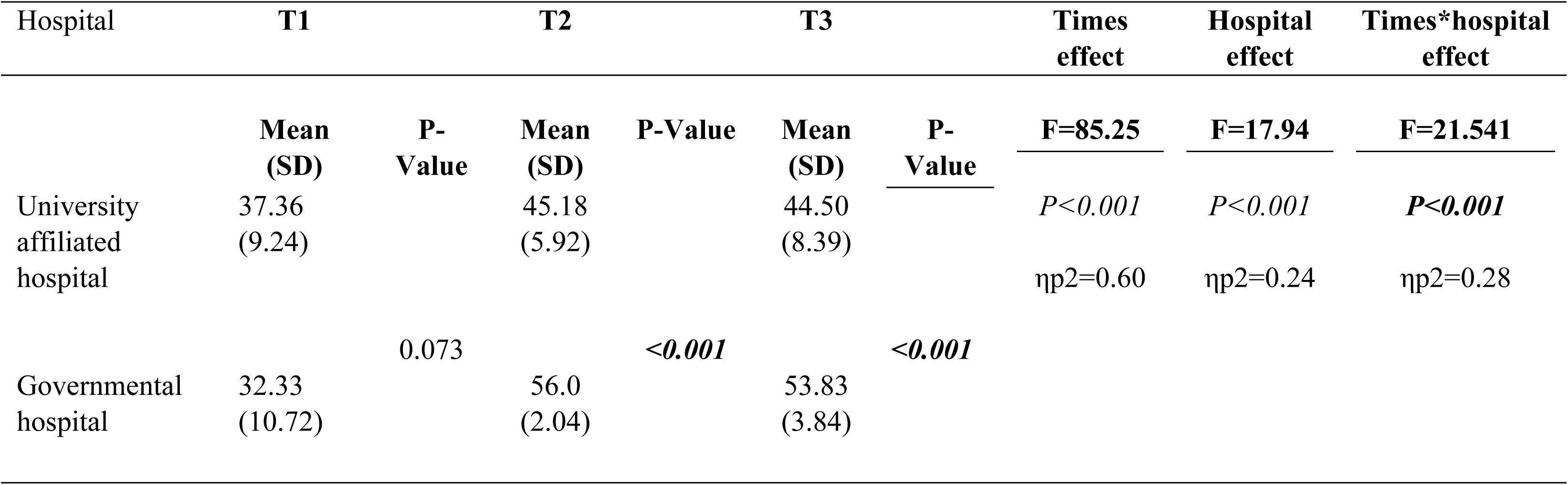
Mean difference in overall knowledge regarding unplanned extubation between the two settings using Split plot ANOVA (n= 58)

Age was negatively correlated with baseline overall knowledge scores (*r* = −0.405, *p* = 0.002), indicating lower UE-related knowledge among older nurses. Knowledge scores were not significantly associated with educational level or years of experience. Post the study intervention the age and overall knowledge negative association was diminished at one week and one month assessment points.

### Effect of the Educational Intervention on Nurses’ Practice

Practice outcomes were assessed only in G-NICU. Nurses’ adherence to UE preventive practices improved significantly over time (*F* = 35.41, *p* < 0.001, ηp² = 0.50). Mean practice scores increased from 9.44 ± 2.63 at baseline to 12.33 ± 1.52 one week after the intervention and further increased to 12.75 ± 1.41 at one-month follow-up (Table 3).

Post hoc comparisons showed significant improvements between baseline and both post-intervention assessments, whereas no significant difference was observed between the one-week post-intervention and one-month follow-up measurements, indicating maintenance of practice gains over time. No demographic characteristics were significantly associated with baseline practice scores and post intervention scores.

## Discussion

The findings obtained from the current study support the effectiveness of educational intervention to enhance neonate nurses’ knowledge of UE and preventive practices. Unlike previous quality-improvement studies that implemented comprehensive UE prevention bundles, and incorporating multidisciplinary engagement, continuous monitoring and staff education, the current study focused exclusively on nurse education. Therefore, the study findings should be viewed as evidence of the effectiveness of education in improving nurses’ knowledge and practice regarding UE prevention rather than reducing UE incidence directly. Given that nurses play a central role in caring for mechanically intubated neonates in the NICU and addressing many modifiable UE risk factors, improvement their knowledge and practice may represent a first step toward reducing UE. Particularly none of the participating nurses had received formal education on UE prevention which highlight a significant educational gap and the need for structured educational programs.

In the current study the baseline UE rates documented in both settings exceeded the commonly recommended benchmark of less than one event per 100 ventilator days and higher than rates reported in United States, Saudi Arabia and Qatar (8,9,11,16,29). The elevated rates observed in the north region in Jordan may be attributed to a number of unit-level factors. These factors including such as limiting cause-analysis reporting of UE events, lack of standardized ETT securement protocols and absence or low sedation which is identified as risk factors for UE (36). Particularly in G-NICU the UE rate was six-fold higher than in U-NICU, suggesting that local organizational practices may substantially influence UE risk. Several differences between the two settings may explain this variation. In U-NICU, ETT taping was typically performed by two nurses using securement approach similar to Pants technique, whereas in G-NICU retaping was performed by one nurse without a clearly defined taping method. Previous studies have shown that the involvement of two healthcare providers during ETT securement and retaping can reduce accidental tube dislodgement (8,18). Furthermore, U-NICU had a limited reporting process of UE events, whereas no formal reporting existed in G-NICU. Event reporting may promote staff awareness and facilitate learning from previous UE events (37).

Among the potential risk factors for UE is parents’ involvement in the care of intubated neonates (37,38). Notably the U-NICU with activated supervised parental involvement in neonatal care showed a sixfold lower UE rate compared to G-NICU which restricted parents’ participation. Although the present study was not aimed to evaluate the association between parental involvement and UE rates, the contribution of parental involvement should be interpreted cautiously due to the existence of many differences between the NICUs. Nevertheless, parental involvement in neonatal care is widely recognized as a cornerstone of family-centered care and is valued by nurses in Jordan for improving communication and supporting infant care (39). (37) et al. (2025) reported that increased staff awareness through regular feedback on UE events contributed to decrease UE rates during skin-to-skin care. This finding suggest that parental involvement can be implemented safely when nurses are adequately educated about UE and its preventive measures. This further emphasizing the importance of nurse education in promoting safe family-centered care in the NICU.

The educational intervention resulted in significant improvements in nurses’ knowledge of UE in both settings as indicated by large effect size ((ηp² = .60). This improvement was evidence after the study intervention and remained significantly higher than baseline at one-month-follow-up. This finding is consistent with adult learning theory, which proposes that educational program aligned with learners’ needs facilitate knowledge acquisition and retention (40). Also, continuing professional development programs perceived as clinically relevant were found to be effective in improving and maintaining nurses’ knowledge (41). In particularly UE-related quality improvement initiatives also, reported enhanced staff awareness and compliance following educational interventions (17,42). Further support is provided by a study conducted in Jordan by Ali et al. (2019) (43), which implemented a four-week educational intervention regarding respiratory distress for neonate nurses. The study indicated a significant improvements nurses’ knowledge with gain maintained at one-month follow-up.

Notably greater improvement observed among nurses in G-NICU which may be attributable to their slightly lower baseline knowledge, providing more opportunity for learning gains. This interpretation is supported by educational research showing that lower baseline performance is often associated with larger subsequent improvement (44).

Although some decline in knowledge scores occurred between the one-week post intervention and one-month follow-up assessment scores remained significantly higher than baseline scores. This suggests successful short-term retention which highlights the value of ongoing reinforcement strategies. For example, regular feedback regarding UE events and associated risk factors has been associated with sustained improvements in staff awareness and reduction in UE rates in the NICU (37).

The current study findings indicated that young nurses had significantly higher baseline UE knowledge compared to old nurses. This finding is consistent with evidence suggesting that younger nurses are more likely to utilize digital educational resources and health informatics technologies thereby increasing their exposure to evidence (45). In the same line Xu et al. (2021) (46) reported that younger healthcare providers demonstrated greater engagement with evidence-based practice and have high informatics-related competencies. Given that the age was not associated with post-intervention knowledge scores in both NICUs, thus the educational programs appeared to be effective across age groups. None of the participating nurses had previous UE formal education, it seems that clinical experience alone is not sufficient to optimize knowledge. This further highlights the need for continuing education and professional development for nurses in such critical units.

Also, in the current study intervention was associated with significant improvements in observed UE preventive practices in G-NICU. Sustained of improvement at one-month follow-up and large effect size (ηp² = .50) suggest that the educational intervention produced meaningful enhancement in nurses’ clinical performance. This finding is consistent with previous UE-quality-improvement initiatives in which staff education served as key components of improved adherence to UE preventive practices in the United State (8,23). Ahn et al. (8) reported greater than 90% compliance with a UE prevention bundle among neonate nurses after the implementation of education initiatives. As well, Caprio et al. (2024) (23) demonstrated improved compliance with evidence-based ETT care following the implementation of a nurse-led UE prevention program in the NICUs.

In our study demographic characteristics were not associated with UE preventive practice scores. this finding is in congruent with a previous study conducted in the NICU in Jordan, demonstrated that enhancing nurses’ evidence-based practices regarding neonatal respiratory distress were not associated with nurse demographic characteristics (43). Collectively, these findings support the role of targeted education and organizational factors more than individual nurse characteristics in promoting evidence-based practice pertained to UE prevention in the NICU in Jordan.

### Implications

the educational gap identifies at baseline and the pivotal role of nurses in preventing UE alongside with limited resources in Jordan emphasize the importance of integrating low-cost strategies such as structured UE education. This may be a practical and sustainable strategy for enhancing the safety of mechanically ventilated neonates in the NICUs. The absence of prior UE-specific training among participants highlights the role of institutions to incorporate UE-prevention education into continuous professional development programs. Both novice and experienced neonate nurses may benefit from targeted education on UE prevention. Such training would support competence development among novice nurses and reinforce evidence-based practice among experienced nurses.

The substantial differences in UE rates observed between the participating settings suggest that organizational practices may significantly influence neonate safety outcomes. Therefore, NICUs should consider developing and implementing standardized UE prevention policies that is cost-effective and culturally sensitive. Such policies including clear ETT securement protocols, standardized UE reporting, identification of high-risk activities or situations, supervised parents’ involvement and staffing recommendations for caring for mechanically intubated neonates.

This study has several limitations. First is the absence of control group which may limit causal inferences regarding the observed improvements in knowledge and practice. Second is that the observation of nursing practice was conducted only in the governmental NICU due to logistical constraints, which may limit the generalizability of the practice-related findings. Third, the one-month follow-up period is not sufficient for evaluation of long-term knowledge retention or sustainability of practice improvements. Thus, future studies should examine long-term effects of educational interventions within this area. Fourth, the use of convenience sampling and recruitment from only two NICUs in the north of the country may affect the generalizability of the current findings. Multicenter studies are recommended to further identifying organizational factors influencing UE prevention and to enhance generalizability of the findings across different NICUs settings. Finally, although the knowledge questionnaire showed excellent validity and reliability, further psychometric evaluation is warranted in a broader neonatal nursing population. Despite these identified limitations, the study provides evidence supporting the effectiveness of a low-cost educational intervention in improving neonate nurses’ knowledge and practice related to UE in the NICU.

## Conclusion

The current quasi-experimental study demonstrated that a structured and clinically relevant educational interventions for one month significantly improved nurses’ knowledge and practice related to UE in the NICU. The gained improvements maintained at one-month follow-up further emphasize the effectiveness of targeted educational strategies. Given the absence of prior UE-specific training among nurses and high baseline UE rates in both settings, thus incorporating UE-focused education into continuing professional development programs would represent a feasible approach to enhance the quality of care for intubated neonates. Further research is needed to evaluate the long-term impact of educational interventions related to UE and its impact on nurses’ knowledge and practice and UE rates in the NICU.

## Data Availability

The data underlying the findings of this study contain sensitive information and are owned by Jordan University of Science and Technology. Ethical and institutional restrictions prevent public deposition of the dataset. The minimal dataset necessary to replicate the study findings will be made available upon reasonable request, subject to approval by Jordan University of Science and Technology and compliance with applicable ethical requirements. Interested researchers may contact the corresponding author to request access.

## Acknowledgment

The authors sincerely thank all neonatal nurses who participated in this study and the administrators and staff of the participating hospitals for their cooperation and support during the implementation of the study and educational intervention. We also gratefully acknowledge the Deanship of Academic Research at Jordan University of Science and Technology for providing financial support for this research.

## Supporting information

**S1 Table. 1 Educational Intervention**. Components of the Educational Intervention for Prevention of Unplanned Extubation in NICUs.

**S2 Table. 2 Knowledge Domains.** Knowledge Domains of Unplanned Extubation (UE) Among NICU Nurses.

**S3 Table. 3 Observational Checklist.** Observational Checklist items for Unplanned Extuabtion the Preventive Practices

